# Large increase in mortality and hospital admissions among young children and the aged due to Influenza and Respiratory Syncytial Virus in Brazil in 2025

**DOI:** 10.64898/2026.08.15.26360470

**Authors:** Emil Kupek

**Affiliations:** Department of Public Health Federal University of Santa Catarina

**Keywords:** Mortality, SARS-CoV-2, Human Influenza, Respiratory Syncytial Virus, Children, Aged, Brazil

## Abstract

**Background:** Mortality and hospital admissions due to Severe Acute Respiratory Infection (SARI) peaked between January and August 2025 in Brazil.

**Methods:** The Brazilian Ministry of Health data on hospital admissions and deaths caused by SARI were compiled by age group (<5, 5-14, 15-49, 50-64, 65+ years) and quarter between January 2023 and June 2025. SARI causes were aggregated into SARS-Cov-2, Influenza, Respiratory Syncytial Virus (RSV), and other viruses (parainfluenza, adenovirus, rhinovirus, bocavirus, metapneumovirus). Multinomial regression was used to impute likely causes of death when these were not laboratory confirmed.

**Results:** In the second quarter of 2025 (2025/2), RSV mortality rate among children <5 years reached 60 per 100,000 – a 43% increase compared with 2024/2. Mortality rate for the joint impact of parainfluenza, adenovirus, rhinovirus, bocavirus, and metapneumovirus in the same age group doubled from 20 to 40 on the same scale in 2025/2 compared to 2024/2. Over the same period, influenza mortality tripled among the aged, whereas mortality due to other respiratory viruses increased less dramatically, except for SARS-CoV-2, which decreased among the aged from 150 to 25 per 100,000 between 2023/1 and 2025/2. Other age groups remained relatively stable over the period. The variation in hospital admissions largely followed that of mortality.

**Conclusions:** While deaths and hospital admissions caused by SARS-CoV-2 declined rapidly since 2023, mortality rates of other respiratory viruses, mainly influenza and RSV, increased significantly among children <5 years and the aged in 2025/2. Public health policies that facilitate vaccine uptake against these infections should be given high priority.

## 1 Introduction

Starting May 2023, the World Health Organisation (WHO) declared the end of the public health emergency caused by the SARS-CoV-2 pandemic, initiated in January 2020 [1]. Despite a drastic reduction in deaths and hospital admissions due to COVID-19, the WHO still considers it an active pandemic because of continued viral activity, its adverse effects, the potential to produce new variants, and to aggravate global health. However, other respiratory pathogens, such as RSV, have caught the attention of both specialists and media, due to increased mortality and hospital admissions [2]. On the other hand, the estimate of 43.7% of Severe Acute Respiratory Infection (SARI) deaths caused by COVID-19 between January and May 2025 [2] casts doubt on a simplistic view that COVID-19 is not a serious health issue in Brazil nowadays.

The present study used commonly applied measures of disease severity – mortality and hospital admissions – to describe quarterly variation of the SARI burden in Brazil and its major components after the COVID-19 pandemic was no longer a public health emergency. Also, these indicators were related to influenza and COVID-19 vaccine coverage. Health policy implications based on the study findings are presented in the discussion.

## 2 Materials and Methods

Data sources on the number of influenza and COVID-19 vaccine doses applied were obtained from the Brazilian Ministry of Health websites *OpenSUS* and *Vacinometro* [3–6]. SARI hospital admissions and causes of death were retrieved from the websites *Dados Abertos* [7] and *SIVEP-Gripe* [8]. Population size by age (0-4, 5-14, 15-49, 50-64, 65 or more years) over the 2023-2025 period was downloaded from the official population statistics web page [9].

The study outcomes were mortality rate (MR) per 100,000 and hospital admission rate (HAR) per 10,000 inhabitants within the aforementioned age groups. The rates were calculated for major causes of SARI: SARS-CoV-2, influenza, RSV, and other respiratory viruses. Case definition was predominantly based on laboratory results, but also included clinical, epidemiological, and medical imaging evidence. In the absence of laboratory confirmation, any piece of this evidence was considered sufficient to define the cause of infection.

Influenza and COVID-19 vaccine coverage were exposure measures of interest. Vaccine coverage was expressed as “vaccines-per-person” (VPP), as the number of vaccine doses applied to each age group in a given calendar year by the population size of that group. Also, influenza vaccine coverage was calculated as the cumulative percentage of vaccinated per month in key target populations: children, the aged, and pregnant women.

An ecological study design with quarterly aggregated data was applied to reduce the effect of notification delay. Such a design is appropriate for showing an overall time trend and including both individual and herd immunity effects.

Statistical analysis first accounted for missing pathogen identification, mainly due to delayed laboratory test results, by estimating the likely pathogen category based on its association with the state of residence, calendar year, month of year, the number of comorbid conditions, patient age, sex, and disease outcome (cure or death). The association was calculated by multinomial regression with complete data and extended (imputed) to the missing data, so each missing pathogen identification was provided with the probabilities of SARS-CoV-2, influenza, RSV, and other respiratory pathogens. The largest of these probabilities defined the most likely cause of hospital admission and/or death and was used in all subsequent analyses.

Time trends for age groups and pathogens were presented graphically. Poisson regression was used to calculate mortality and hospital admission rates with 95% confidence intervals (CI).

Stata software, version 17.0, was used for statistical procedures and data preparation.

## 3 Results

This section presents key findings on missing data, followed by graphical summaries of quarterly variation in MR and HAR. As most of the Poisson regression 95% confidence limits were within ±10% of their estimates, these limits were omitted from the graphs for clarity, but can be obtained in tabular form on request.

Multinomial regression estimated that 12.33%, 23.56%, 27.72%, and 36.40% of missing pathogen identification were likely influenza, RSV, SARS-CoV-2, and other respiratory viruses, respectively. It is remarkable that about 49% of 883,248 health records lacked pathogen specification months after being notified to epidemiological surveillance of SARI cases. Imputation results pointed to an underestimation of 67% of influenza cases, 92% of COVID-19, 50% of RSV, and 55% of other SARI viruses. As epidemiological surveillance bulletins and disease burden calculated thereof in Brazil are based almost exclusively on confirmed cases, the likely impact of these infections on population health is often underestimated.

In the 2^nd^ quarter (spring season) of 2025, RSV mortality rate among children under 5 years reached 60 per 100,000 – almost a 50% increase compared with the same quarter in the previous year (Figure 1). Over the same period, mortality due to parainfluenza, adenovirus, rhinovirus, bocavirus, and metapneumovirus together approximately doubled, from 20 to 40 per 100,000.

**Figure 1.**
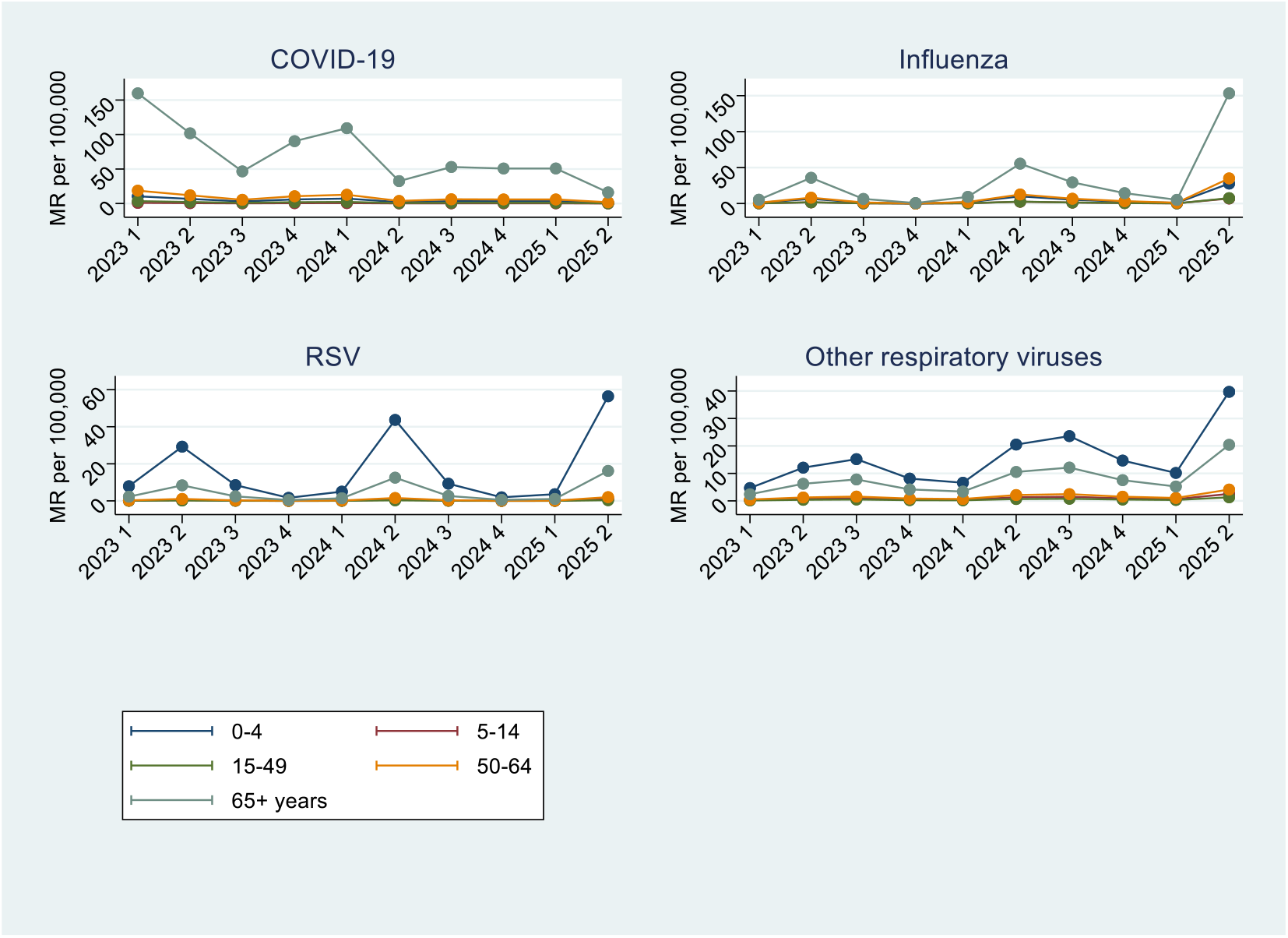
Mortality rate (MR) for major causes of Severe Acute Respiratory Infections in Brazil between January 2023 and June 2025, by age

Among the aged (65+ years), influenza MR increased from 50 per 100,000 in the 2^nd^ quarter of 2024 to 150 in the same quarter of 2025. Over the same period, mortality rates for RSV and other respiratory viruses except SARS-CoV-2 also increased, whereas the latter dropped from 150 to 25 per 100.000 between the 1^st^ quarter of 2023 and the 2^nd^ quarter of 2025. Other age groups showed relatively small mortality variation over the period.

HAR for SARI by age followed the time trends similar to those of MR trends (Figure 2). The SARS-CoV-2 HAR per 10,000 among the aged dropped from 18 to <5, whereas the influenza HAR rose from 5 in the 2^nd^ quarter of 2024 to 15 in the same quarter of 2025. Among children under five years, the HAR doubled in the 2^nd^ quarter of 2025 due to influenza, so 0.1% of this age group were admitted to the hospital (Figure 2). Between the 2^nd^ quarters of 2024 and 2025, the HAR per 10,000 increased from 43 to 70 due to RSV and from 18 to 25 due to other viruses.

**Figure 2.**
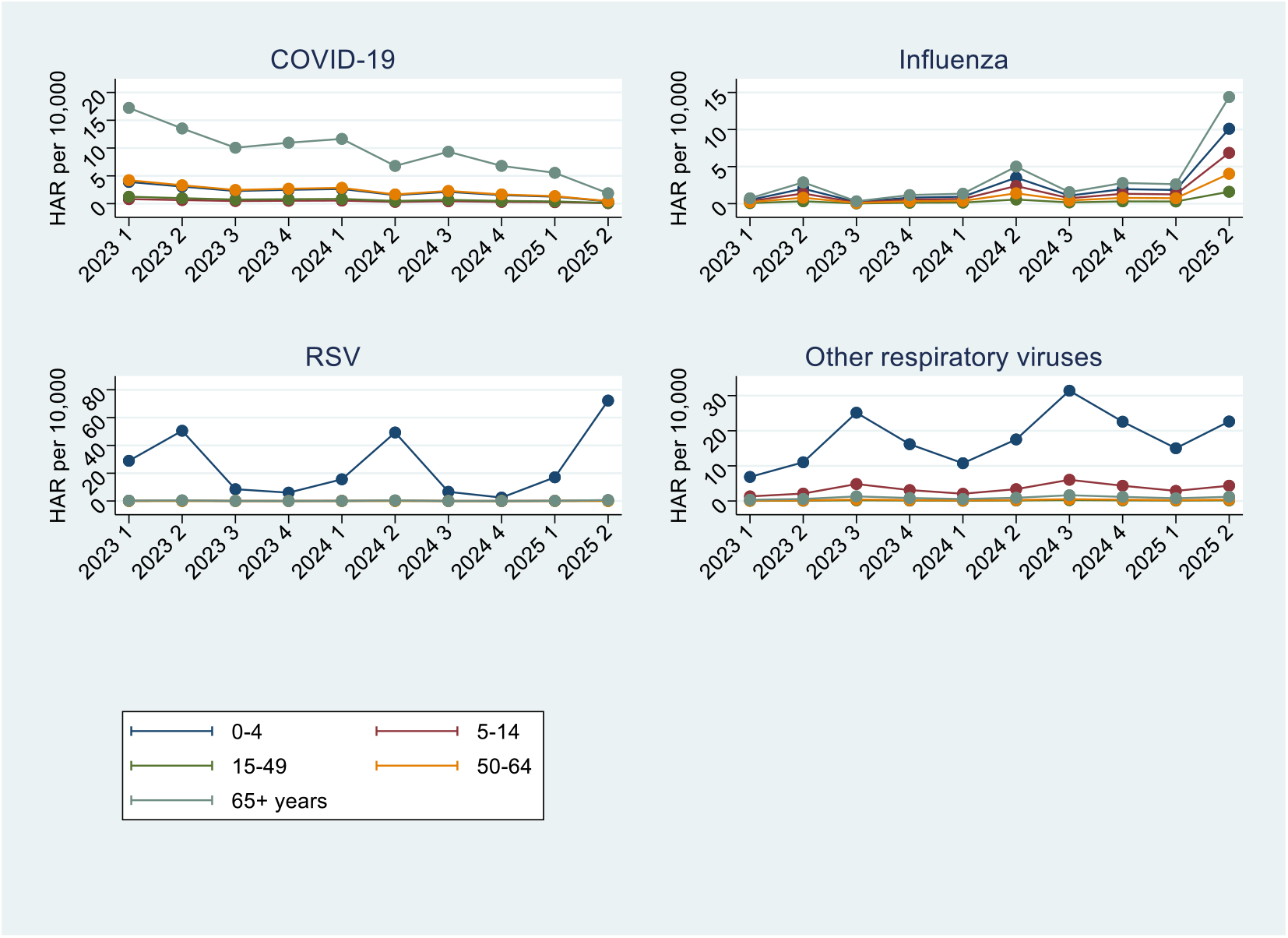
Hospital admission rates (HAR) for major causes of Severe Acute Respiratory Infections in Brazil between January 2023 and June 2025, by age

Seasonal increases in influenza mortality in the second quarters were preceded by low vaccine coverage for this infection among children and the aged (Figure 3). Following the increase in mortality, vaccine coverage of these age groups improved within the same quarter, with accentuated reduction in influenza mortality in subsequent quarters. Cumulative influenza vaccine coverage by month of 2025 (Figure 4) shows that neither pregnant women nor the aged reached 50% vaccine coverage in the 2^nd^ quarter of 2025, when influenza mortality peaked. By the end of the quarter, children <6 years had slightly better coverage at 60%.

**Figure 3.**
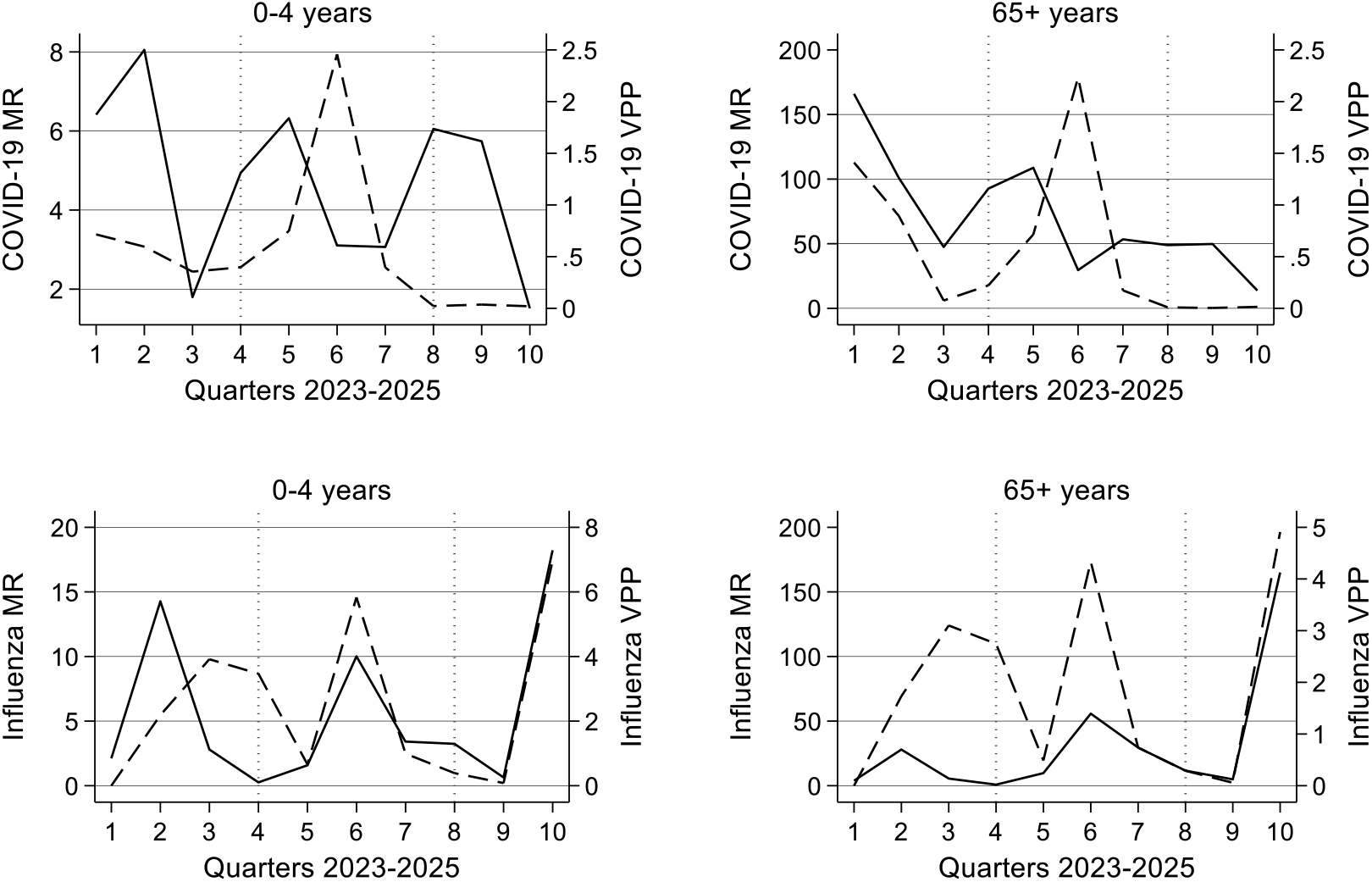
COVID-19 and influenza mortality rates per 100,000 (continuous line) and the number of vaccine doses applied per person (dotted line) in Brazil for the most affected age groups between January 2023 and June 2025

**Figure 4.**
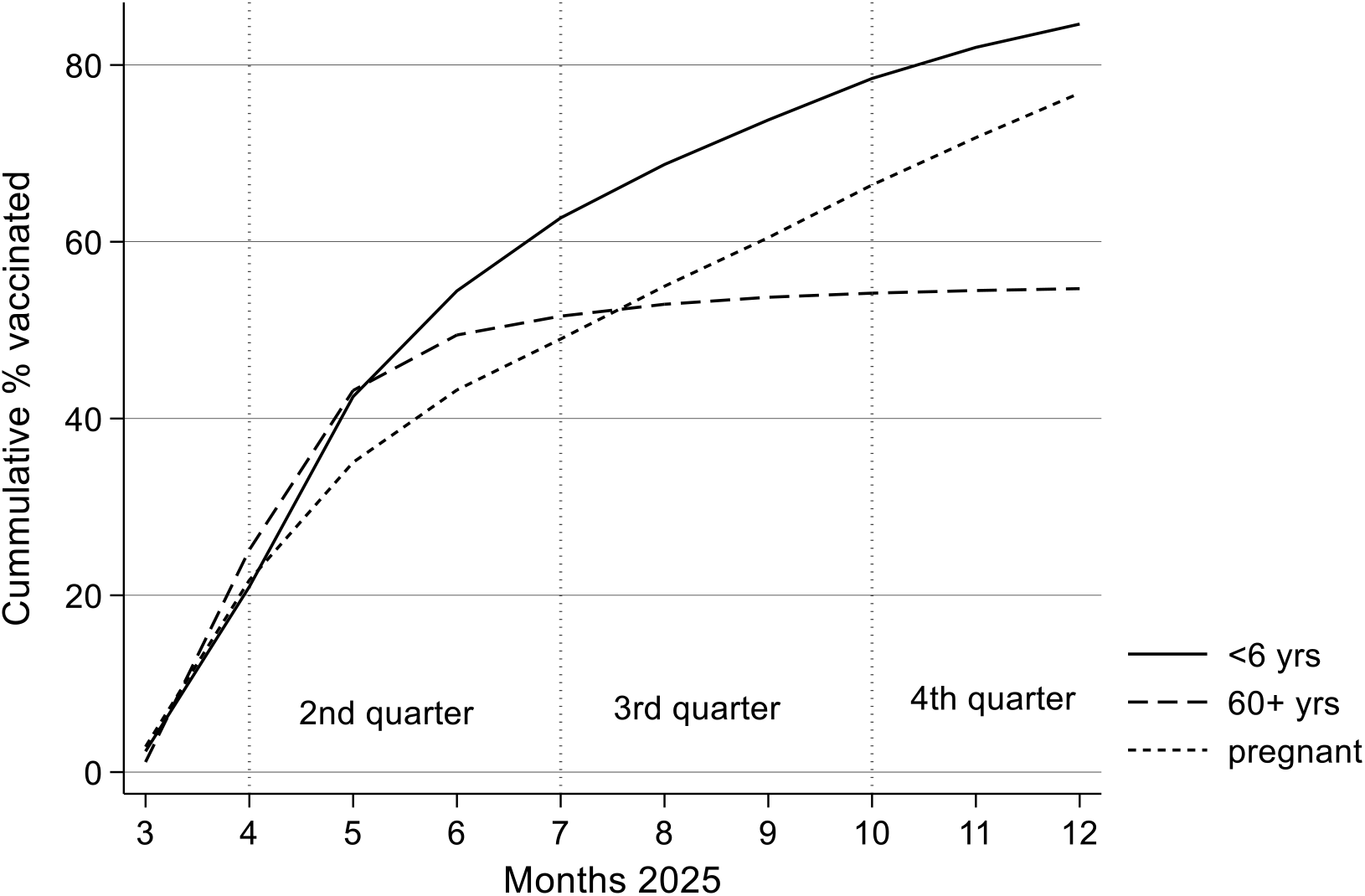
Cumulative influenza vaccine coverage (%) over the months of 2025 for key target groups in Brazil: children between 6 months and <6 years, people of 60 years and older, and pregnant women

In 2024, the COVID-19 vaccine uptake accelerated in parallel with the rise in COVID-19 MR (Figure 3). In the first half of 2025, both COVID-19 vaccine coverage and its MR decreased to almost zero.

## 4 Discussion

This is the first epidemiological report on a large increase in SARI mortality and hospital admissions in Brazilian children and the aged in the 2^nd^ quarter of 2025, which identified major pathogens and accounted for reporting delay in laboratory test results. Other age groups’ mortality and hospital admission rates were considerably less variable between January 2023 and June 2025.

For comparison’s sake, in 2023 pneumonia-related MR per 100,000 in Brazil was 12 among children under the age of five years and 292 among people 65 years or older, with general population MR of 40 [10]. In 2025, the latter value passed 100 [11] and drew media attention [12]. In light of the present study results, this increase could have been partially driven by complications of respiratory viral infections reported in the study.

A rising contribution of respiratory viruses, such as RSV, parainfluenza, adenovirus, rhinovirus, bocavirus, and metapneumovirus, has already been reported in Brazil [13]. The present study found that these viruses doubled MRs to 40 per 100,000 in this age group in the 2^nd^ quarter of 2025, thus reaching the aforementioned pneumonia-related mortality. During the same period, SARS-CoV-2 MR was about half that value. Taken together, these data highlight the need to expand child immunisation against circulating pneumococcal strains and RSV, in addition to already available universal vaccination for influenza and SARS-CoV-2, within the Brazilian national vaccination program.

A Brazilian cohort with over two-thirds of children under five years found that influenza vaccine was over 70% effective against hospital admission or death in this age group and 46.3% effective in the general population [14]. It also prevented 2 in 3 intensive care unit admissions. Lower vaccine coverage and effectiveness in older people, found in the present study (Figure 3), may be influenced by more frequent comorbidities in older age. It is worth noting that influenza vaccination intensified in the same quarters in which an expressive increase in mortality was observed among the aged, suggesting that the vaccination was largely reactive to this increase, therefore at least a month too late. Cumulative influenza vaccine coverage <50% among the aged and pregnant women, and <60% among children, points to insufficient immunisation of key population segments the vaccination aimed at.

SARS-CoV-2 pandemic altered seasonal influenza patterns in the southern hemisphere [15], making it more difficult to plan and execute a mass vaccination. Also, in Brazil, the 2^nd^ quarter of 2025 was colder than average, especially in the southern and southeastern regions, which may have contributed to enhanced transmission of respiratory infections.

Between February 2020 and February 2023, SARS-CoV-2 was a major cause of SARI hospital admissions in Brazil and led to 6.5% of all SARI deaths with known pathogen [16]. Over the same period, influenza contributed 2.3%, whereas adenovirus, VSR, and other respiratory viruses together caused 7% of the aforementioned deaths - more than COVID-19. In other words, mortality attributed to respiratory viruses other than SARS-CoV-2 was already high during the pandemic, then jumped to an even higher level once the pandemic was no longer a public health emergency.

Among study limitations, it is worth keeping in mind those inherent to secondary data and ecological study design, such as reporting delay, memory error, missing information, and the use of a proxy measure of vaccine coverage (VPP). However, despite its limitations, VPP has already been applied in a similar context [17]. The study strengths include large, regularly updated data on key outcomes, exposure and control variables, and the use of standard statistical techniques to account for missing values. Furthermore, the magnitude of seasonal peaks in mortality and hospital admissions enhances the validity of key findings.

Study findings underscore the importance of precise vaccination timing. It is relevant not only in Brazil but also in global public health policies as respiratory viruses circulate rapidly worldwide. While vaccination is widely recognised as the most cost-effective public health intervention to prevent these infections, its implementation is often challenging due to vaccine hesitancy, financial and logistical difficulties of scaling up vaccine production and distribution, especially in large countries with different climate zones. The One Health perspective is well suited to address these issues and evaluate vaccine effectiveness in reducing disease burden not only at the individual level but also at the country and wider regional level.

Health policy implications of the study results should be given high priority and tailored to tackle country-specific barriers to better vaccine coverage. In many low-income countries, the cost of vaccines against influenza, COVID-19, and RSV may be prohibitive, so international aid programs should be considered, despite the current political climate moving in the opposite direction. In Brazil, an upper-middle-income country with sufficient financial resources to produce or buy these vaccines and adequate primary care services to implement the vaccination, adjustments to improve access to health services would be very helpful. The latter have working hours that coincide with those of commerce and industry, and therefore are difficult to attend by working-class people. More flexible working hours during the week, offering vaccines in public places with large circulation (shopping malls, busy metro or bus stations) and specific target groups (e.g. children in school or pre-school institutions) has been applied sporadically, but needs to be scaled up. Also, combating disinformation that fuels vaccine hesitancy is of utmost importance, albeit challenging. Despite state media efforts to encourage free-of-charge vaccine uptake in Brazilian primary care facilities, reaching out through social networks has proved much more difficult. With over half of the population relying on these as a primary information source, health promotion of vaccines must innovate in this area to get its message through.

## 5 Conclusions

In Brazil, a significant reduction in COVID-19 mortality and hospital admission rates occurred after SARS-CoV-2 was no longer a public health emergency. However, influenza, RSV, and other respiratory viruses causing SARI showed the opposite trend, with steep peaks in the second quarter (spring season), mainly among children under five years and the aged. SARI burden is often underestimated with currently available surveillance unless missing data is accounted for.

## Data Availability

All data produced are available online at the URL indicated in the references.

## Availability of data and materials

The datasets supporting the conclusions of this article are available in the following repositories:

OpenDataSus: https://opendatasus.saude.gov.br/dataset/srag-2021-a-023

OpenDataSus: https://opendatasus.saude.gov.br/dataset/covid-19-vacinacao/resource/301983f2-aa50-4977-8fec-cfab0806cb0b

Vacinômetro COVID-19: https://infoms.saude.gov.br/extensions/SEIDIGI_DEMAS_Vacina_C19/SEIDIGI_DEMAS_Vacina_C19.html

Dados Abertos. SRAG 2021 a 2025 - Banco de Dados de Síndrome Respiratória Aguda Grave - incluindo dados da COVID-19: https://dados.gov.br/dados/conjuntos-dados/srag-2021-e-2022

Dados/InfoGripe: https://gitlab.fiocruz.br/marcelo.gomes/infogripe//blob/master/Dados/InfoGripe/obitos_semanais_fx_etaria_virus_sem_filtro_febre.csv

